# Feasibility of adjuvant self-administered intravaginal 5-fluorouracil cream following primary treatment of cervical intraepithelial neoplasia grade 2 or 3 among women living with HIV in Kenya: study protocol for a pilot trial

**DOI:** 10.1101/2023.12.13.23299916

**Authors:** Chemtai Mungo, Elizabeth Bukusi, Grace E Kirkland, Cirillus Ogollah, Gershon Rota, Jackton Omoto, Lisa Rahangdale

## Abstract

**Background:** Women living with HIV (WLWH), the majority of whom live in low- and middle-income countries (LMICs), are at higher risk of cervical precancer, known as cervical intraepithelial neoplasia (CIN), and are up to six times more likely to get cervical cancer. Current CIN treatment methods, primarily ablation or excision, have high treatment failure rates among WLWH, up to 30% for CIN grade 2 or 3 (CIN2/3) at 24 months following ablation. Without strong follow-up many WLWH with treatment failure are at risk of developing invasive cervical cancer, highlighting the urgent need for improved CIN treatment methods. Prior studies in high-income countries (HICs) have demonstrated that 5-Fluorouracil (5-FU) cream, an antimetabolite drug that is easily accessible in LMICs, can be used intravaginally as adjuvant therapy following primary CIN2/3 treatment in WLWH to reduce CIN2/3 recurrence. While the safety, acceptability, and efficacy of self-administered 5-FU for cervical precancer treatment has been demonstrated in HICs, it has not been studied among WLWH in LMICs who bear the greatest burden of cervical cancer.

**Methods:** We are conducting a Phase I pilot study investigating the feasibility of using 5-FU cream as an adjuvant, self-administered intravaginal therapy following cervical intraepithelial neoplasia grade 2/3 (CIN2/3) treatment among WLWH in Kenya (ClinicalTrials.gov NCT05362955). Twelve participants will be enrolled in this single-arm study. Participants will self-administer 2g of 5% 5-FU cream intravaginally every other week for eight applications. The primary objective is to determine safety, defined as the type, frequency, and severity of adverse events (AEs) using a standardized grading scale. The secondary objectives are uptake, tolerability, adherence, and acceptability.

**Results:** There are no results at this time as this is an ongoing study.

**Discussion:** To achieve the World Health Organization (WHO) 90/70/90 global cervical cancer elimination goals, which include 90% of women with cervical precancer adequately treated by 2030, it is essential to employ innovative and resource-appropriate strategies to improve cervical precancer treatment among WLWH. The use of 5-FU as adjuvant therapy following current screen & treat programs may be a feasible and scalable strategy to optimize outcomes in this high-risk group. This clinical trial will provide important feasibility data to inform future randomized efficacy trials in LMICs.

**Trial registration:** ClinicalTrials.gov identifier: NCT05362955

**Key messages regarding feasibility:** *What uncertainties existed regarding feasibility?:* There are no uncertainties to report yet as the study is ongoing.

*What are the key feasibility findings?:* There are no key feasibility findings as the study is ongoing.

*What are the implications of the feasibility findings for the design of the main study?:* Feasibility findings from this trial that will inform the design of the future study include safety findings - frequency and severity of adverse events, which may necessitate a different dosing schedule for the main study, adherence findings - if low levels of adherence to self-administered therapies are established, the authors will work on designing strategies to improve adherence prior to moving to a larger study. Finally, acceptability findings including the levels of male partner support of the treatment will inform future study efforts.

## Background

Although cervical cancer is preventable, in 2018, an estimated 570,000 new cases occurred, with low- and middle-income countries (LMICs) accounting for 85 percent of incident cases and 90 percent of deaths.^1^ Cervical cancer is an AIDS-defining malignancy and women living with HIV (WLWH), the majority of whom reside in LMICs, are at significantly increased risk.^2^ Compared to HIV-negative women, WLWH have increased incidence and persistence of human papillomavirus (HPV),^3^ have a shorter time from HPV infection to development of precancerous lesions,^4^ and have a six to eightfold increased risk of developing invasive cancer,^5,6^ making prevention efforts among them an urgent priority. With increasing access to antiretroviral therapy (ART), WLWH in LMICs have prolonged life expectancy, increasing their vulnerability to death from cervical cancer.^7^ Unfortunately, increasing access to ART, and hence improved immune status has not brought about a decrease in cervical cancer incidence in LMICs.^4^ Lack of widespread screening programs coupled with a lack of accessible and effective treatment of precancerous lesions in LMICs accounts for the observed disproportionate burden of cervical cancer.^8,9^

Recent efforts have focused on developing alternative, feasible, and resource appropriate cervical cancer prevention tools for implementation in LMICs, including among WLWH.^10,11,12^ The World Health Organization (WHO) recommends HPV testing for cervical cancer screening in LMICs, followed by treatment without histopathologic verification in a single-visit approach, to reduce loss-to-follow up.^13^ Recent innovations, including use of point-of-care HPV self-collection, hold significant promise for expanding the reach of screening programs in LMICs. However, to achieve the WHO’s global strategy on cervical cancer elimination, including 70% of women screened, and 90% of those with a positive result adequately treated by 2030,^14^ significant attention is warranted on improving treatment of precancerous lesions among WLWH living in LMICs. Available precancer treatment methods in LMICs are primarily ablation or excision - primarily Loop Electrosurgical Excision Procedure (LEEP). Ablative methods which use frozen gas (cryotherapy) or heat (thermal ablation) to destroy precancerous cells, are less technical than excision, can be performed by nurses, and are the mainstay of precancer treatment outside of referral facilities, including among WLWH.^13,15^ Thermal ablation, approved by the WHO in 2019, is most readily available and scalable due to its lower cost and availability of portable, battery-powered, reusable devices that can be used in mobile campaigns and rural clinics.^16^ However, among WLWH, both ablative and excisional treatments are limited by high rates of cervical intraepithelial neoplasia grade 2 and 3 (CIN2/3) recurrence. Randomized trials in South Africa and Kenya demonstrate 18-19% and 27-30% CIN2/3 recurrence following LEEP and cryotherapy among WLWH, respectively.^3,17^ The high rates of CIN2/3 recurrence are driven in part by high rates of high-risk human papillomavirus (HPV) persistence following precancer treatment among WLWH.^18^ This highlights an important limitation even with universal access to precancer treatment among WLWH, and most concerning in LMICs where attendance at one-year post-treatment follow-up visits (where recurrence can be identified) is as low as 24% due to weak healthcare care systems.^19^ This calls for studies on innovative, accessible, adjuvant treatments, including topical therapies,^20–26^ immunotherapy,^27^ therapeutic vaccinations,^28–30^ and other novel approaches for treating precancerous lesions among WLWH.^31^ One such promising therapy is self-administered topical 5-fluorouracil (5-FU).

Topical 5-FU has been studied as a primary or adjuvant treatment for cervical precancerous lesions with promising results.^21,24–26,32^ 5-FU is an antimetabolite and cytotoxic agent which acts through inhibition of DNA and RNA synthesis.^33,34^ 5-FU is currently FDA approved for the treatment of a variety of malignancies, including superficial basal cell carcinoma.^35^ Observational and randomized trials have demonstrated tolerability and efficacy of topical 5-FU for treatment of genital warts,^36^ vulvar,^37^ anal,^38^ and cervical^25,26^ precancers. In a prospective randomized trial of off-label intravaginal self-administered 5% 5-FU versus observation among US HIV-uninfected women with CIN2, primary treatment with 5-FU was associated with an 84% regression of disease, compared to 52% in the observation group (RR 1.62; 95% CI 1.10 – 2.56) at six months.^26^ Importantly, in this trial, topical 5-FU dosed every two weeks was well tolerated, with no grade 3 or 4 adverse events (AEs), or symptoms that caused interference with usual activities.^26^ Additionally, 86% of participants reported satisfaction with use of topical 5-FU.^26^ In another US-based randomized trial of self-administered 5% 5-FU versus observation for adjuvant treatment in HIV-infected women following standard (excisional or ablative) treatment for CIN2/3, 5-FU was associated with improved outcome, with CIN 2/3 recurrence identified in 8% of participants in the 5-FU arm compared to 31% of participants in the observation arm (p=0.014) at 18 months.^25^ Similarly, in this study with biweekly only 2% of women reported mild AEs with treatment, and no grade 3 or 4 AEs were reported.^25^

These data suggest that 5-FU, a readily available generic therapy in LMICs on the WHO list of recommended medications^39^, may be a novel, safe and accessible topical, self-administered adjuvant therapy to prevent CIN2/3 recurrence following primary treatment in WLWH in LMICs. However, no studies have evaluated the feasibility of repurposing topical 5FU for this indication in LMICs where the burden of cervical cancer is greatest. To fill address this research gap and inform future efficacy and implementation trials, we designed a Phase I pilot study to evaluate the safety, uptake, tolerability, adherence, and acceptability of adjuvant, self-administered intravaginal 5-FU among WLWH in Kenya who have had primary CIN2/3 treatment.

## Methods

### Study Objectives

#### Primary Objective: Safety

The primary objective of safety will be evaluated as type, frequency, severity of adverse events among study participants. Adverse events will be evaluated using a standardized, validated, grading scale developed by the National Institute of Health, Division of AIDS for Female Genital Grading for Microbicide Studies.^40^ Adverse events will be graded as grade 0- none, grade 1- mild, grade 2- moderate, grade 3- severe, grade 4- potentially life threatening. Adverse events will be elicited on history, reviewed from symptom diaries, and evaluated on pelvic exam, including evaluating for inflammation, erythema, erosions or other changes. The at-risk period for safety will begin at study Week 1 (first week of 5-FU use) and will continue through study Week 20 (4 weeks after last 5-FU use), or the last attended safety evaluation in the case of premature study exit.

### Secondary Objectives

1. Uptake will be defined as proportion of eligible screened participants who agree to participate in the trial.
2. Tolerability will be defined as the defined as the proportion of participants who are unable or unwilling to apply at least 4 of 8 doses (50%) of 5% 5-FU cream due adverse events, burden of self-application, etc.
3. Adherence will be evaluated by assessing the number of participants who are confirmed to use 75% or more of the 5-FU applications (i.e., at least 6 of 8 doses). A participant will be classified as adherent to the specific 5- FU dose if on adherence evaluation, 2 of 3 of the methods of evaluating adherence support 5-FU application of that particular dose.
4. Acceptability will be evaluated using a questionnaire as well as through in-depth interviews among study participants at the end of the trial.

### Study design

This is a single arm, nonrandomized, interventional pilot trial among 12 women living with HIV, age 18 – 49 years in Kisumu, Kenya. Participants who have biopsy-confirmed CIN2/3 and have undergone primary treatment with standard of care ablation or excisional procedures will be eligible for enrollment. The study intervention is use of 2 grams of self-administered 5% 5-FU cream intravaginally, once nightly, every other week for 8 applications. Outcomes including safety, tolerance, and adherence will be ascertained longitudinally during the study by trained clinicians. The protocol was developed in line with the Standard Protocol Items: Recommendations for Interventional Trials (SPIRIT) 2013 checklist guidelines (see Supplementary materials).^41^

### Study setting

The study will take place at Lumumba Sub County Hospital, a referral facility in Kisumu County, Kenya, in East Africa. Like many LMICs, Kenya faces a high burden of both HIV and cervical cancer. The national HIV prevalence in Kenya is approximately 4.5%.^42^ In comparison, Kisumu County and surrounding areas have disproportionately higher HIV prevalence of 17.5%,^43^ over three times the national prevalence. According to the 2018 Kenya Cancer Guidelines, cervical cancer contributes 5,250 (12.9%) of the new cancer cases annually and 3, 286 (11.8%) of all annual cancer deaths.^44^ Cervical cancer screening is offered in Kenya primary using visual inspection with acetic acid, and precancer treatment is primarily ablation with a few referral facilities offering excisional procedures.^45^

### Eligibility and recruitment

Participants will be recruited from outpatient clinics offering cervical cancer screening and cervical precancer treatment in Kisumu County as well as neighboring counties. Study staff will provide educational talks about the study, and interested women undergoing cervical precancer treatment will be evaluated for eligibility. Potential participants will be screened to determine if they meet the study eligibility criteria (Table 1). Participants meeting eligibility criteria will sequentially be enrolled until the planned sample size is met.

**Table 1:**
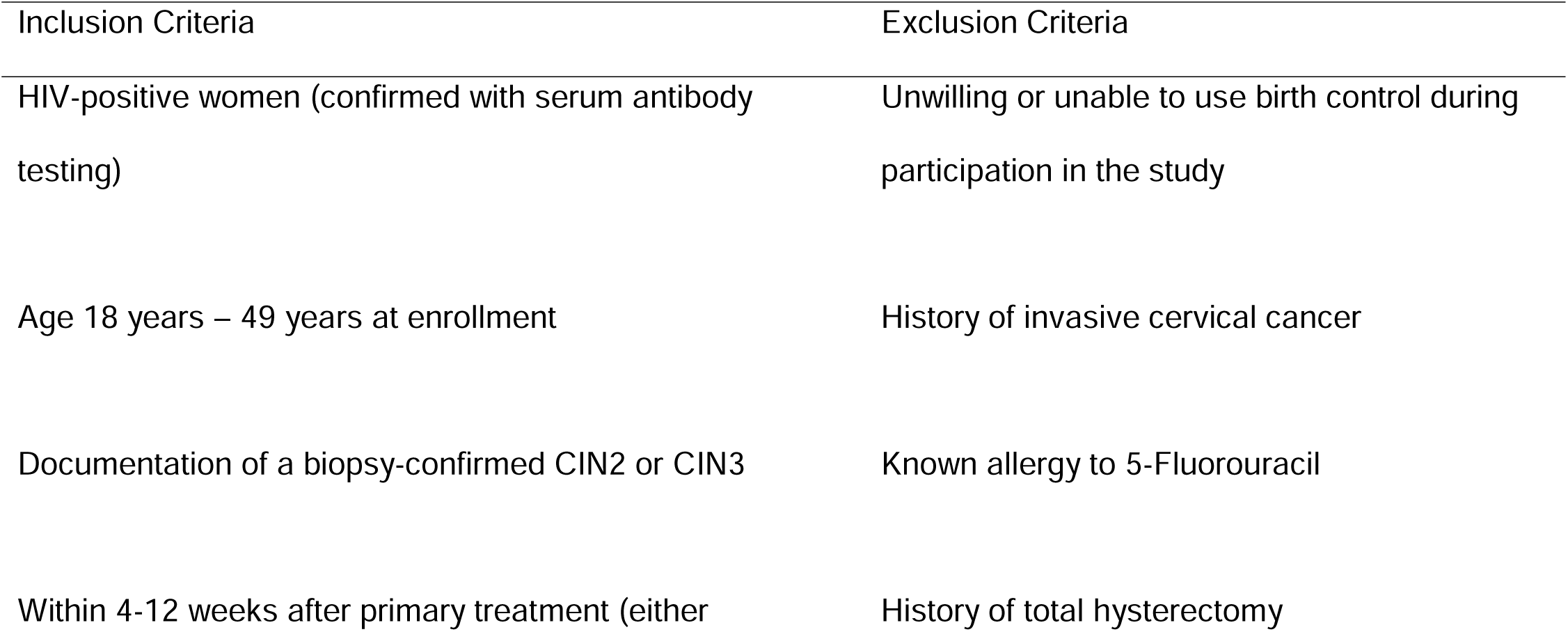
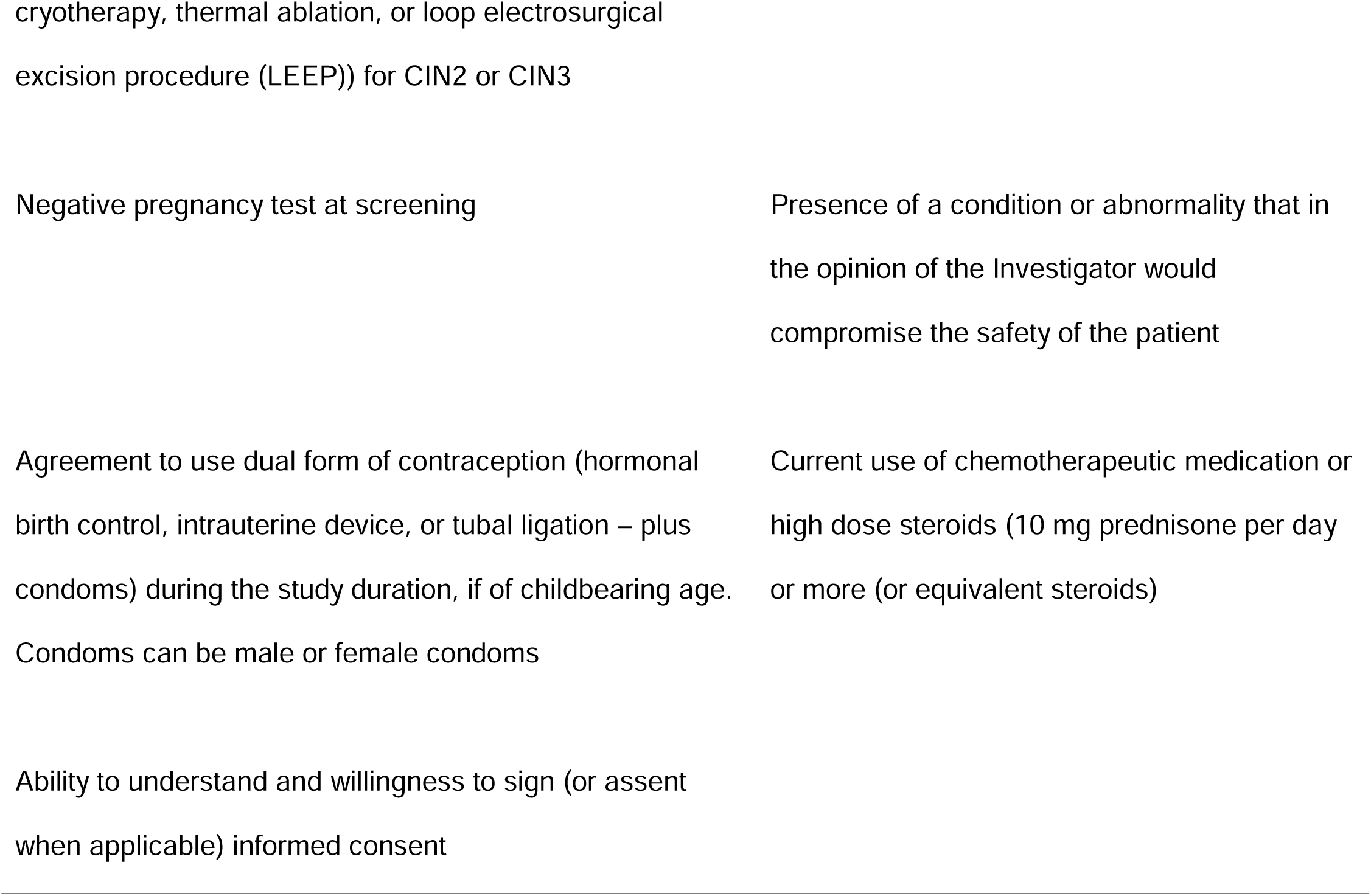
Inclusion and exclusion criteria for study participants.

### Study procedures by visit

#### Week 0: Screening Visit(s)

The study procedures are illustrated in Figure 1. Potential study participants will be assessed for eligibility over a series of screening visits. Informed consent will be obtained before any study procedures are performed. During screening, a pregnancy test will be performed, and the HIV status will be confirmed using a rapid HIV test and a second confirmatory method. A complete medical history will be obtained, including verification of a history of biopsy-confirmed CIN2/3, being within 4-12 weeks of primary treatment and review of concomitant medications. A physical exam will be performed, including a pelvic and colposcopy exam, to ensure appropriate healing of the cervix following primary treatment. Specimens for HPV testing, gonorrhea, and chlamydia testing will be obtained, as well as serum specimens for CD4 count testing. Participants diagnosed with any sexually transmitted infections will be treated before enrollment. Detailed protocol counseling will be performed, including 5FU use duration and frequency, expectations for abstinence around 5FU use, frequency of study visits and duration of the study. Participants will be given the option to bring in their male partners or other support persons whom they feel may need to be part of the counseling process as part of the consenting process. Complete locator information will be obtained, including two phone numbers and a home location to enable tracking if needed and acceptable to the study participant.

#### Week 1: Enrollment Visit and 5FU use education

Following ascertainment of eligibility, participants will attend an enrollment visit within 28 days of the screening visit. At this visit, pregnancy testing will be repeated, and an interval medical history will be obtained, including the use of any new medications. Detailed protocol counseling will be conducted by a study nurse, including detailed counseling on how to self-administer 5FU at home (Box 1), which will be done at night once every two weeks for 8 applications. Prior to use, participants will be instructed to wash their hands with soap and water and apply a petroleum-based emollient, such as plain Vaseline to the labial area prior to 5FU use for barrier protection. The 5FU application will be performed with a graded applicator with a clearly demarcated 2g mark, which will be provided to applicants. Participants will be encouraged to place a tampon in the vagina overnight following 5FU use to keep the cream at the cervix, which will be removed in the morning and discarded. On rising in the morning after 5FU use, participants will be instructed to shower or rinse, and clean the labia with soap and water, and apply more barrier-protection to the labia, continuing for 1-3 days. Participants will also be advised to use a panty liner for 1-3 days after each 5FU use, as needed, for comfort, which will be provided alongside other supplies. Participants will be counseled on the need to abstain from sex for at least two days after each time they use 5FU, as well as the need to consistently use condoms throughout the study period, alongside another form on contraception.

##### Box 1.

###### Instructions for Intravaginal 5-Fluorouracil (5FU) Application

1. Use 5% 5FU cream provided
2. Wash hands with soap and water
3. Apply petroleum-based emollient such as plain Vaseline to the labial area prior to 5FU use
4. Fill the applicator provided with 2g of 5FU, gently insert the applicator into the vagina until resistance is met, and dispense the cream by pushing the applicator to the end
5. Place a tampon in the vagina overnight to keep the cream in the vagina
6. Place the applicator in the resealable bag provided to bring to the clinic on your next visit
7. In the morning, remove and discard the tampon away from children. Rinse or take a shower to clean the area well. Apply more petroleum-based emollient to the labia and use pantyliners for 2-3 days after 5FU use to protect the skin.
8. Avoid sexual intercourse for at least two days after applying 5FU and use condoms on other days
9. Use 5FU once every other week, on the same day, for eight applications total

All women treated with topical 5-Fluorouracil were provided these standardized instructions. The instructions were reviewed with each participant by a study nurse or doctor, and a comprehension checklist was used to confirm understanding.

At the enrollment visit, 5FU self-administration teaching will be done by a study nurse or doctor, using pictorial aids as well as the use of a pelvic model to demonstrate insertion of the applicator up to the cervix and dispensing of the drug. During the demonstration in the clinic, toothpaste will be used instead of 5FU cream to demonstrate the loading of the drug onto the applicator and correct application. Participants will be advised to use the 5FU cream at night before they go to sleep, on the days they will use it, and to pick one day of the week to use 5FU every other week for eight applications. As part of 5FU self-administration teaching, participants will be taught how to attach the 5FU tube to the provided applicator and how to load and measure 2g of 5FU onto the graded applicator to ensure correct dosing. Using a pelvic model, a demonstration of 5FU administration will be performed by a study nurse, including how to gently insert the applicator intravaginally until resistance is met (indicating the cervix), how to deploy the cream by advancing the applicator until all the cream is dispensed, and removal of the applicator which will be placed in a resealable plastic bag to be returned to the study clinic at the next appointment. Following this demonstration, all study participants will have multiple opportunities to practice all steps of 5FU self-administration, including connecting the tube to the applicator, correctly measuring 2g, advancing the applicator on the pelvic model until they meet resistance, deploying the cream, and playing a tampon on the pelvic model. Participants will practice self-administration as many times as is necessary to become competent. Education and counseling will be done in the participant’s preferred languages, including Swahili or Dholuo. A competency checklist (Supplement 1), which will be performed by a different provider, will be used to ensure each participant is competent in all aspects of 5FU self-administration before the study drug is dispensed. The study drug, 5% 5FU cream, will be provided in a 20g tube, which participants will take home. All participants will be given the option of self-administration of the first dose of 5FU in the clinic under the observation of a nurse, if they prefer, to increase their confidence in self-administration and receive feedback. At the enrollment visit, all participants will be provided a study bag containing the study drug, applicators, condoms, tampons, panty liners, resealable bags to place used applicators and petroleum emollient. Additionally, literate participants will be provided a study diary, onto which they will record the day they use the 5FU cream, as well as record any adverse events they have. Participants will be instructed to bring the study bag to each clinic visit, including the used applicators placed in a separate resealable bag, the study drug, unused applicators, and the study diary. A study phone number will be provided to participants to call or send a text message with questions. Menstruating participants will be instructed to call the study clinic if they have their menstrual period on the day they are scheduled to use 5FU, in which case use will be delayed until the end of the menses.

#### Weeks 1, 3, 5, 7, 9, 11, 13, 15: 5FU self-administration at home

Participants will self-administer 5FU at home on one day of the week, on weeks 1, 3, 5, 7, 9, 11, 13, 15, based on the instructions provided. On Week 1 (enrollment week), participants will have the option of self-administration of 5FU in the study clinic under the observation of a study nurse.

#### Weeks 2, 4, 6, 8, 10, 12, 14: Safety and adherence assessments

Participants will return to the study clinic on the intervening weeks (weeks 2, 4, 6, 8, 10, 12, 14, 16) in between 5FU use for safety and adherence assessments. As part of the safety evaluation, adverse events (AEs) will be elicited and reviewed. Written adverse events on participant study diaries will be assessed, AEs will be elicited on history and a pelvic exam and colposcopy will be performed to evaluate for pelvic AEs. At each safety visit, a urine pregnancy test will be performed to rule out pregnancy. At these events, adherence assessment will be performed to ensure correct use of the study drug, which will include a review of the study diary for when participants used 5FU, as well as a self-report of 5FU use on the prior week. During these weeks, the study team will maintain regular telephone contact with study participants on phone and text messages to support adherence.

#### Week 16

Participants will return to the study clinic on Week 16, after using the final (eight) dose of self-administered 5FU at home on Week 15. At this visit, similar safety and adherence assessments as weeks 2-14 will be performed, including a pelvic exam and colposcopy for magnified visualization of the cervix. During the pelvic exam, a specimen for HPV testing will be collected. Participants will bring their remaining 5FU drug which will be collected and discarded. An acceptability questionnaire will be administered, and an in-depth interview performed as part of acceptability assessment.

#### Week 20: Final follow-up and study exit

All participants will return in Week 20 for a final study visit, during which time any delayed adverse events will be elicited or evaluated. A pelvic exam will be done only based on history (as needed). The participant will then exit the study.

#### Strategies to promote adherence to 5FU self-administration

Several strategies will be utilized to promote adherence to 5FU self-administration, including in-depth education at study enrollment, use of text message or phone call reminders, and biweekly study visits during the dosing phase. At enrollment, detailed education on 5FU self-administration, as outlined above, will be performed with each participant. A comprehension checklist will be used to ensure all participants understand all steps of 5FU self-administration and are able to replicate the steps in the study clinic using a pelvic model. Once ready, participants will be given the option to self-administer the first dose of 5FU in the clinic under guidance of a study nurse, if they prefer. To support adherence to the scheduled once every other week dosing, participants will be encouraged to select one day of the week which they will use the 5FU and use the study drug consistently on the same day. A study calendar will be provided to literate participants to support adherence. Finally, all participants will receive a weekly text message or phone call from the study team reminding them of their planned 5FU use (every other week), as well as reviewing any adverse events they have. During the biweekly clinic visits during the dosing phase, adherence will be reinforced during counselling sessions by the study team.

#### Safety evaluations

Safety evaluation to evaluate for adverse events will be done using the standardized Division of AIDS Table for Grading the Adverse Events for Female Microbicide Trials.^40^ Evaluations will include a pelvic exam, which will be performed by a study clinician, during which genital tissues will be evaluated for drug-related adverse outcomes, including inflammation, erythema, ulceration, or other adverse events based on the standardized grading scale (grade 0 – normal, grade 1 – mild, grade 2 – moderate, grade 3 – severe, grade 4- potentially life-threatening). Adverse events (AEs) duration (in days), severity/grade, outcome, treatment, and relation to study drug will be recorded in the participant study records. grade 3 or 4 AEs (possibly, probably, or definitely related to the study drug) will be reported to the DSMC within 5 business days of the PI becoming aware. Participants found to have any grade 3 or 4 AEs will be instructed to stop study drug use immediately and will not resume until the Data Safety and Monitoring Committee decides that the AE is not related to the study drug.

#### Adherence evaluation

Three methods will be used to evaluate whether participants used the self-administered 5FU doses at home per the study protocol; 1) self-report, 2) weighing of study tubes at each visit, 3) ultraviolet (UV)-light assessment of returned applicators. For self-report, participants will record in their study diary the day they used the 5FU and will bring the diary to the clinic visits for review by a study clinician. Participants will also bring the 5FU tube dispensed at baseline during each study visit, which will be weighted biweekly to document reduction in weight consistent with expected 5FU use the week prior. Lastly, participants will be asked to return the used vaginal applicators – placed in a resealable bag that will be provided. The applicators will be inspected under ultraviolet light for evidence of intravaginal insertion which has been validated in vaginal microbicide trials.^46,47^

#### Participant retention plan

Once a participant is enrolled, study staff will make every effort to retain the participant for the protocol-specified duration of follow-up. Participants will be enrolled in the study only if they demonstrate understanding of the study and study procedures, including length of follow-up and what is expected of them during study participation, which will be assessed through the comprehensive assessment conducted during the informed consent process. At enrollment, detailed contact information will be obtained, including phone number(s) and residence information in case a participant needs to be traced after a missing visit. Study staff will communicate with participants regularly including via phone calls or text messages, as part of promoting adherence to 5FU self-administration, as well as to promote retention. To facilitate adherence and mitigate participant’s expenses associated with study visits, participants will be reimbursed for each study visit, as outlined in the informed consent form and approved by the institutional review boards. If a participant misses a study visit, a study team member will make every effort to contact the participant by phone or physical tracing except when the participant withdraws consent, or the visit is not feasible due to unplanned location. Only participants who plan to stay within the study locale during the study duration will be enrolled.

### Statistical Considerations

#### Sample size

Twelve participants will be enrolled in the study, and up to 2 participants will be replaced if an enrolled participant drops off the study before week 8.

#### Sample size Justification

Prior studies on the use of intravaginal 5-FU studies at this dosage and frequency have not reported any grade 3 or 4 AEs.^13,14^ As this study will use the same dosing/frequency schedule, we expect no grade 3 or 4 AEs. If there are no any grade 3 or 4 AEs, as expected, then based on an exact binomial confidence interval and a sample size of 12, the upper limit of the 95% confidence interval of the AE rate will be 26.5%, that is, we will be able to confidently rule out severe AEs greater than 26.5% in this population.^30^ Additionally, if ≥ 33% (4 or more participants) experience a dose-limiting toxicity (DLT) – defined as a severe AE (grade 3 or 4), we will not proceed with the planned Phase II study. Dr. Rahangdale’s trial of 60 HIV-negative women did not result in any AEs that were grade 2 or higher.^14^ Therefore, we do not expect to observe a toxicity rate of greater than 30% in our study. The probability of observing DLT in 4 or more participants for toxicity rates no greater than 30% are listed in Table 2. We expect grade 1 and 2 AEs may be more common in this trial. Table 3 gives the anticipated precision for different possible grade 1 and 2 AEs for the planned sample size.

**Table 2:** Projected dose-limited toxicity rate.

| Projected dose-limited toxicity (DLT) rate | Probability of observing DLT in $\geq 33\%$ of participants |
| --- | --- |
| ≤ 5% | < 1% |
| 10% | 2.6% |
| 20% | 20.5% |
| 30% | 50.7% |

**Table 3:**
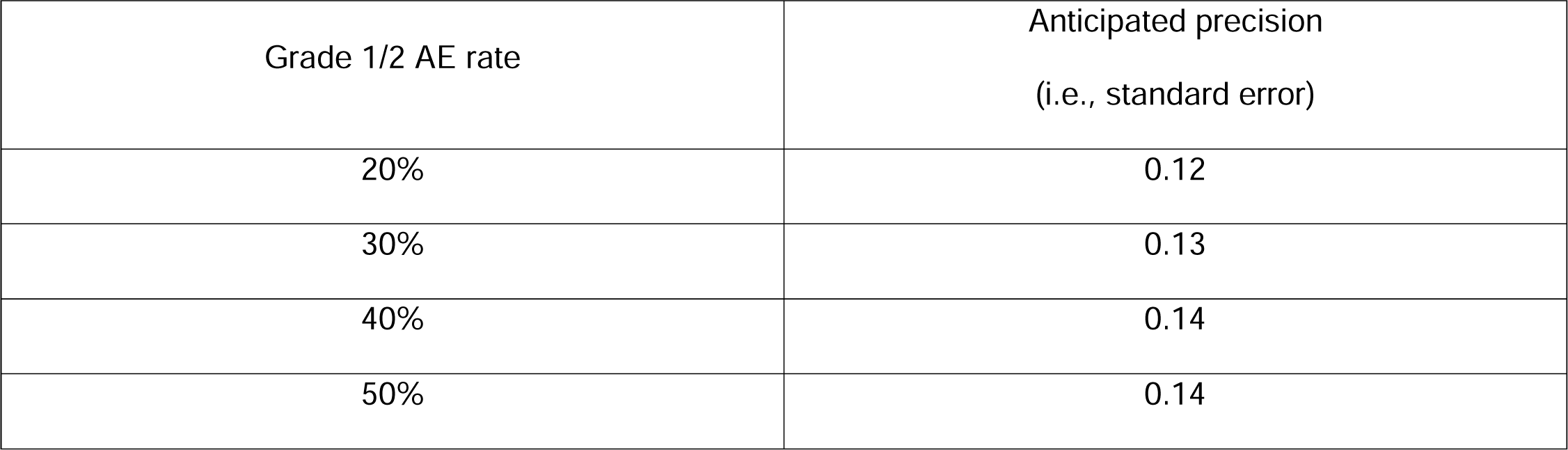
Anticipated precision for grade 1 and 2 adverse events.

| Grade 1/2 AE rate | Anticipated precision<br>(i.e., standard error) |
| --- | --- |
| 20% | 0.12 |
| 30% | 0.13 |
| 40% | 0.14 |
| 50% | 0.14 |

### Data Analysis

For the primary endpoint of safety, we will report the type and frequency, and severity of reported adverse events (AEs), including exact 95% confidence intervals. The at-risk period for safety will begin at study Week 1 (first week of 5-FU use) and will continue through study Week 20 (4 weeks after last 5-FU use), or the last attended safety evaluation in the case of premature study exit. For secondary endpoints, uptake will be evaluated as the proportion of eligible screened participants who agree to participate in the trial. For tolerability, we will report the proportion of participants unable to apply at least 50% of the doses, with exact 95% confidence intervals. For adherence, we will report the proportion (and 95% confidence interval) of participants confirmed to use at least 6 of the 8 planned 5FU doses, with corresponding 95% confidence intervals. A participant will be classified as adherent to the specific 5-FU dose if on adherence evaluation, at least 2 of the 3 methods of evaluating adherence support 5-FU application. Safety, tolerability and adherence data will be reported for all enrolled participants including early dropouts or withdrawals. Acceptability will be evaluated including means and standard deviation for responses to questions graded on a Likert scale on the acceptability questionnaire, and proportions and 95% confidence intervals for yes/no questions. In-depth interviews (IDIs) to elicit participants experiences with 5FU self-administration, including challenges associated with the treatment will be done with each study participant. In-depth interviews will be conducted in local languages in a private room, audio recorded, transcribed and coded using thematic analysis. Interviewers will elicit participants experiences with side effects, partners support of abstinence requirements, and comfort with the intervention.

### Data safety and monitoring

The University of North Carolina (UNC) Chapel Hill Lineberger Comprehensive Cancer Center (LCCC) Data and Safety Monitoring Committee (DSMC) will review the study on a regular (quarterly to annually) basis, with the frequency of review based on risk and complexity as determined by the UNC Protocol Review Committee. The DSMC is comprised of faculty affiliate members of the Lineberger Comprehensive Cancer Center, with backgrounds in Oncology, Biostatistics, Pharmacy and Public Health, and is independent of the sponsor and has no competing interests. The UNC PI will be responsible for submitting the following information to the DSMC for review: 1) safety and accrual data including the number of subjects treated; 2) significant developments reported in the literature that may affect the safety of subjects or the ethics of the study; 3) preliminary response data; and 4) summaries of team meetings that have occurred since the last report. Findings of the DSMC review will be disseminated by memo to the UNC Principal Investigator, the UNC Protocol Review Committee, the any of the regulatory bodies charged with the safety of human subjects and the integrity of data including, but not limited to, the Kenya and UNC Institutional Review Boards (IRBs), the oversight of Office of Human Research Ethics (OHRE) Biomedical IRB, the Oncology Protocol Review Committee (PRC) or the North Carolina TraCS Institute Data and Safety Monitoring Board (DSMB).

## Discussion

This pilot trial will evaluate the safety and feasibility of repurposing 5FU cream, a generically available drug in most LMICs that is on the WHO List of Essential Medications, as a self-administered, adjuvant therapy following primary CIN2/3 treatment among WLWH who face 25% - 30% treatment failure rate following both ablation and excision. The trial will provide data on safety of self-administered 5FU in this high-risk group, which, if demonstrated, can pave the way for efficacy studies and provide a potentially affordable and scalable strategy to improve cervical precancer treatment among WLHW in LMICs. The study will evaluate other feasibility outcomes, including uptake of the intervention among eligible participants, adherence to self-administered 5FU using both self-report and objective measures, tolerance to the study drug regimen, and acceptability among both women and their male partners, through survey and qualitative evaluation of study participants. The study regimen, 2 g of 5% 5FU cream, used every other week for 8 doses, has previously been demonstrated to be safe and tolerable among both HIV-positive^48^ and HIV-negative^49^ women in HICs, when used as a self-administered therapy. If a similar safety profile can be established, with evidence of adherence and acceptability, topical, self-administered 5FU can be evaluated in randomized trials among WLWH in LMICs to improve CIN2/3 treatment outcomes following current ablative or excisional therapies.

### Current Status

The study opened for recruitment in May 2023 and is currently recruiting. The study is expected to finish follow-up in May 2024.

## Supporting information

Supplemental File - 5FU Self-Administration Checklist

## Data Availability

All data produced in the present study are available upon reasonable request to the authors

## Trial registration

The trial is registered under U.S Clinical trial registry (clinicaltrials.gov, NCT05362955).

## List of abbreviations

LMIC: low- and middle-income countries
LCCC: Lineberger Comprehensive Cancer Center
DSMC: Data safety and monitoring committee
HIV: human immunodeficiency virus
WLWH: women living with HIV
HPV: human papillomavirus
ART: antiretroviral therapy
WHO: World Health Organization
LEEP: Loop Electrosurgical Excision Procedure
CIN2/3: cervical intraepithelial neoplasia grade 2 and 3
5-FU: 5-fluorouracil
AEs: adverse events
DLT: dose limiting toxicity
IDIs: in-depth interviews

## Declarations

### Ethics approval and consent to participate

This clinical trial has full ethics review board approval from the University of North Carolina Chapel Hill, the Kenya Medical Research Institute. Written informed consent will be obtained from all study participants.

### Consent for publication

Not applicable.

### Availability of data and materials

Not applicable.

### Authors’ contributions

CM and LR conceived and designed the study, providing subject matter expertise and overseeing all aspects of protocol development. EB offered guidance on subject matter, ethical considerations, collaborative management, and study implementation. JO & COO provided guidance on protocol development and implementation and will lead protocol implementation in country. EB offered guidance on subject matter, ethical considerations, collaborative management, and study implementation. LR (Co-Investigator) provided expertise on protocol implementation, data analysis, interpretation, and Phase II grant writing. JO (Co-Investigator) contributed subject matter expertise, study design, protocol implementation, and capacity building for providers. CO (Co-Investigator) played a crucial role in managing field operations, while GR (Co-Investigator & Study Pharmacist) oversaw study medicine implementation, adverse effects reporting, and regulatory compliance. All authors, in their respective roles, contributed to study and manuscript preparation and have collectively approved the final manuscript.

## Acknowledgements

Not applicable.

