## Supplemental File - 5FU Self-Administration Checklist for "Feasibility of adjuvant self-administered intravaginal 5-fluorouracil cream following primary treatment of cervical intraepithelial neoplasia grade 2 or 3 among women living with HIV in Kenya: study protocol for a pilot trial"

5FU Administration Checklist

This checklist checks the participants understanding of 5FU administration. It should be administered before consent is signed and during weekly visits to reinforce proper 5FU use.

| **Question/Statement** | **Required points of comprehension** | 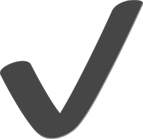 |
| --- | --- | --- |
| How is 5FU applied? | Self-application using an intravaginal applicator |  |
| What amount of 5FU is applied | 2grams |  |
| How often is 5FU administered | Once every other week/ Every 2 weeks before bedtime on the same day |  |
| When can 5FU administration be delayed? | If day of administration coincides with menses, in this case, no two doses should be administered **within 7 days of** each other |  |
| Can the participant demonstrate using the model how to use 5FU? | Participant should be able to appropriately demonstrate how to apply 5FU:   1. Wash hands before handling 2. Connect intravaginal connector to 5FU cream 3. Dispense 2g 4. Lie on back with knees drawn towards chest 5. Insert applicator into the vagina, advance until resistance is felt signifying the position of the Cervix 6. Push plunger to dispense the 5FU cream 7. Withdraw applicator and place it inside self- sealing bags 8. Wash hands and securely store study drug |  |
| Following 5FU administration what should the participant do | Abstain for **48 hours,** on resuming sexual activity continue using condoms + another contraceptive  method |  |
| What should the participant do when they have side effects | 1. Anticipate common side effects such as discharge and mild irritation- i.e for discharge use pantyliners. 2. For side effects affecting normal daily activities, call a study number or seek medical care at the nearest facility. Inform study staff every time this happens |  |
