## Supplemental Figure 1 for "Feasibility of adjuvant self-administered intravaginal 5-fluorouracil cream following primary treatment of cervical intraepithelial neoplasia grade 2 or 3 among women living with HIV in Kenya: study protocol for a pilot trial"

**Figure 1: Summary of Study Procedures**

**Screening Visit**

Review eligibility criteria, obtain informed consent

Clinic visit

*Review medical history*

*Baseline clinical/demographic characteristics*

*Pelvic and colposcopy exam > HPV sample collection*

*Urine collection > gonorrhea and chlamydia, pregnancy test*

*Blood draw > HIV ***, CD4 testing*

$$\downarrow$$

**Week 1: Enrollment Visit**

Teaching and demonstration of intravaginal 5FU application

Clinic visit

*Informed consent and eligibility criteria review*

*Interval medical history*

*Urine pregnancy test*

*Dispense 5-FU and symptom diary, tampons, condoms*

$$\downarrow$$

**Week 1, 3, 5, 7, 9, 11, 13, 15**

Home application of 5-FU

$$\downarrow$$

**Week 2, 4, 6, 8, 10, 12, 14**

Adverse events review and adherence assessment

Clinic Visit

*Symptom diary and adverse events review*

*Adherence assessment*

*Review of concomitant medication*

*Urine pregnancy test*

*Pelvic exam and colposcopy evaluation*

$$\downarrow$$

**Week 16**

Adverse events review and adherence assessment

Clinic Visit

*Symptom diary and adverse events review*

*Adherence assessment*

*Review of concomitant medication*

*Urine pregnancy test*

*Pelvic exam and colposcopy evaluation*

*HPV sample collection*

$$\downarrow$$

**Week 20**

Final follow-up

Clinic Visit

*Adverse event evaluation*

*Pelvic exam as needed*

**Weekly Contact**

Telephone reminders of study timelines
